# A Multi-Schedule Machine Learning Pipeline for Medicare Reimbursement Change Prediction and Operational Risk Stratification

**DOI:** 10.64898/2026.07.22.26358736

**Authors:** Ishan H. Patel, Alejandro Leyva, Muhammad Khalid Khan Niazi

## Abstract

FeePredict is a three-stage random forest machine learning framework to simultaneously predict whether Medicare reimbursement rates for specific procedures will change, in which direction they will change, and by how much. FeePredict was applied to the four major Medicare fee schedules: the Clinical Laboratory Fee Schedule (CLFS), the Physician Fee Schedule (PFS), the Ambulance Fee Schedule (AFS), and the Durable Medical Equipment, Prosthetics, Orthotics, and Supplies (DMEPOS) fee schedule. Each of these fee schedules contains publicly available data from the Centers for Medicare & Medicaid Services (CMS) for the years 2024, 2025, and 2026, with the number of procedures represented in the data ranging from 3,264 to 2,952,842 observations.

FeePredict utilizes lag-1 feature engineering and train-only preprocessing steps to ensure that there is no data leakage into the model. Chronological out-of-time validation was performed on three of the four fee schedules to determine the generalizability of the model over time. FeePredict significantly outperformed the assumption that there would be no changes to Medicare reimbursement rates for procedures (*p <* 0.001), achieving concordance indices between 0.815 and 0.998, and reducing the mean absolute error for predicting changes to reimbursement rates by 29% to 85%.

Permutation testing of the model with shuffled reimbursement rate labels indicates that there is no evidence of data leakage (AUC values: 0.467-0.515). The model achieved concordance indices of 0.854 and 0.972 for the CLFS and DMEPOS fee schedules, respectively, outside of its training period, but performed less well outside of its training period for the PFS, indicating that it generalizes less well to changes to the Medicare policy regime that existed after its training period. Overall, though, these results indicate that it is possible to accurately predict whether Medicare reimbursement rates for medical procedures will change using only data from the historical versions of those fee schedules.

## 1 Introduction

The fee schedules established by Medicare are used to reimburse approximately $900 billion in annual fee-for-service healthcare services, yet health systems lack tools to anticipate the changes to these fee schedules issued by the Centers for Medicare & Medicaid Services (CMS) each year. For fee-for-service healthcare services four main fee schedules exist that must be accounted for in relation to the changes that are to be made to these fee schedules by the CMS: the Clinical Laboratory Fee Schedule (CLFS), Physician Fee Schedule (PFS), Ambulance Fee Schedule (AFS), and the Durable Medical Equipment, Prosthetics, Orthotics, & Supplies Schedule (DMEPOS). Each of these fee schedules has its own rate-setting update mechanism implemented by CMS each year, allowing each to use its own estimation model to forecast the changes likely to occur. Consequently, the development of viable models for predicting the changes to these fee schedules is of great interest to the financial and operational planners within each health system, as they are forced to model for these changes within their annual budget and contract negotiations with providers well in advance of the time that the CMS makes the actual changes to the fee schedules. Currently, many of these providers review the proposed CMS rules for the upcoming year to estimate rate changes for each fee schedule. However, this process is often time-intensive and relies on general fee schedules rather than code-level, prediction-based estimates for each. Thus, there are no systematic tools for health systems to effectively anticipate changes to these fee schedules before CMS publishes them.

In predictive financial healthcare, machine learning methods have been applied to predict individual patients’ healthcare costs [6, 3], plan-payment risk adjustment [7, 8, 9], and high-cost patient identification [11, 10, 18]. Deep learning approaches have further extended these methods to temporal claims data [16], and automated billing-code assignment from clinical text has reached considerable maturity [12, 20]. However, these existing models share a fundamental limitation: they predict the cost of care delivered or the coding of services rendered, not the reimbursement rate that CMS will publish for a given procedure code. The reimbursement literature itself is strongest on validating how rates are set [13, 15] and explaining how providers respond once rates change [14], but does not produce prospective forecasts. To our knowledge, no prior work has developed a generalizable, leakage-aware framework for forecasting Medicare reimbursement-rate changes at the HCPCS code level across multiple fee schedules simultaneously.

Each fee schedule poses different challenges for predicting its code change rates, including varying rate change prevalence, different rate-setting mechanisms, and varying available data for each schedule. In response to this area and challenge in the healthcare world, we have developed FeePredict, a three-stage random forest machine learning model, with each stage targeting unique challenges for each fee schedule. Each of the three stages of FeePredict can be used to effectively predict whether a code will change within a fee schedule, the type of change (increase or decrease) to that code, and the magnitude of the change in healthcare services. Each of the three FeePredict stages has been applied in some form to all four fee schedules. The experiments performed for each fee schedule included model cross-validation, lagged features to prevent leakage, label-shuffle sanity tests, and tests of out-of-time generalizability (except for AFS, which is explained in 2.2).

This topic is covered in this paper, beginning with Section 2, which is dedicated to the Materials and Methods for the FeePredict model and tests. This section fully describes the methods used to train and test the FeePredict model for each fee schedule. Following this section are the results of each of these tests for each of the fee schedules. In Section 3 (Results), the results reveal that each FeePredict model significantly outperformed the baseline of zero change across the data sets for all four fee schedules. In interpreting these results in Section 4 (Discussion), it was found that the models exhibited strong generalizability beyond the training data across all schedules (except AFS, which is explained in 2.2) and revealed some issues with the PFS Fee Schedule. Furthermore, there are discussions of other past work in the field and its relationship to this present work. Finally, Section 5 covers the limitations regarding the data used to train the models. The last section of this paper, Section 6, discusses the conclusions that can be drawn from the studies of the FeePredict model and its capabilities.

## 2 Materials and Methods

### 2.1 Data Sources and Study Cohort

The data was sourced and downloaded directly from the publicly available fee schedules files posted by the Centers for Medicare and Medicaid Services (CMS) on the CMS website (cms.gov). No patient data, claims data, or other protected health information was accessed in the creation of this study. The data consists of four different fee schedules published by Medicare.

Across all four schedules, each row represents one entity in one time period, where the entity and period are defined to match the level at which CMS publishes rates for that schedule. Row counts therefore vary substantially across schedules because the dimensionality of each schedule differs. CLFS and PFS publish a single national rate per HCPCS code per quarter, while AFS rates vary by contractor and DMEPOS rates vary by state, rural-area flag, and equipment modifier. We preserved this granularity in our entity definitions to allow the model to learn rate dynamics at the level CMS actually sets them. The columns in every schedule contain the entity identifiers, the time period, the reimbursement rate, and engineered lag and history features derived from prior periods.

#### Clinical Laboratory Fee Schedule (CLFS)

The CLFS tracks the reimbursement for clinical diagnostic laboratory tests. The CLFS consists of 9 files published quarterly over the 2024-2026 period, with the entity defined at the HCPCS code level. This yielded 16,638 code-quarter observations across 2,053 unique HCPCS codes. After removing rows with no lag features or history, 14,585 observations were compiled into a data set ready for input to the model. The characterization of the CLFS indicates that there were 1.4% changes in the rates for the laboratory tests, and that 97.8% of those changes were increases.

#### Physician Fee Schedule (PFS)

The PFS tracks the reimbursement for physician and non-physician practitioner services. As with the other fee schedules, 9 PFS files were published quarterly over the 2024-2026 period, with the entity defined at the HCPCS code level. The PFS yielded 152,202 code-quarter observations across 16,996 unique HCPCS codes. After removing rows with no lag features or history, 134,530 observations were compiled into a data set ready for modeling. The characterization of the PFS indicates that there are 24.1% changes in the rates for physicians’ and non-physician practitioners’ services, comprising 36,672 change events. These changes are mainly due to adjustments to the annual conversion factor applied to the fee schedule each January, and changes to the RVU values for each procedure code every quarter. Additionally, 45.6% of the changes were increases in the rates for those practitioners, while 54.4% were decreases. There is no class imbalance in the direction of changes in the practitioners’ rates, indicating that the PFS is the only fee schedule with a genuine bidirectional classification problem.

#### Ambulance Fee Schedule (AFS)

The AFS determines the reimbursement for ambulance services. The AFS makes available fee-schedule files published annually for 2024-2026, with the entity defined at the HCPCS-contractor level. Locality was not included in the entity because each of the 55 contractors is responsible for multiple localities within the country; including the locality would cause lag features to be incorrectly assigned to the ambulance services. The files contain 3,264 HCPCS-contractor-year observations across 12 unique HCPCS codes distributed among 55 contractors, yielding 660 unique HCPCS-contractor entities over the three-year window. The characterization of the AFS indicates a 74.9% change in the ambulance rate. More specifically, 70.9% of those changes were rate increases, while 29.1% were rate decreases. Because there were so few unique ambulance procedures coded by HCPCS, a leave-one-group-out cross-validation algorithm was used, with groups defined by HCPCS code.

#### Durable Medical Equipment, Prosthetics, Orthotics, and Supplies (DME-POS)

The DMEPOS schedule tracks the reimbursement for durable medical equipment and related supplies. The DMEPOS fee schedule consists of 9 files published quarterly between 2024 and 2026, with the entity defined at the HCPCS-modifier1-modifier2-state-rural-flag level. Beyond the HCPCS code for each product, the modifiers indicate whether it was a new purchase (NU), a rental (RR), or used equipment (UE), so including the modifier within the entity helps ensure that pricing information for different classes of DMEPOS products is represented in the data. The schedule includes 6,651,500 entity-quarter observations across 2,104 unique HCPCS codes and 372,590 unique entities. After data processing to remove rows with no lag features and invalid entities, 2,952,842 observations were compiled and ready for model input. The schedule shows 14.9% of DMEPOS codes changed during the period, characterized by 99.7% rate increases and 0.3% decreases. Additionally, 5% of the DMEPOS codes were assigned to the rural area of the United States (RURALFLAG = R). For these 5% of codes, there were 1,045,123 instances where the rate was permanently 0, indicating a lack of coverage for those durable products in rural areas of the country.

Summary dataset characteristics across all four schedules are presented in Table 1.

**Table 1:** Dataset characteristics across all four Medicare fee schedules. Rows represent model-ready observations after removal of first-period rows with no lag history.

| Characteristic | CLFS | PFS | AFS | DMEPOS |
| --- | --- | --- | --- | --- |
| *Data Scale* |  |  |  |  |
| Raw rows | 16,638 | 152,202 | 3,264 | 6,651,500 |
| Model-ready rows | 14,585 | 134,530 | 3,264 | 2,952,842 |
| Unique HCPCS codes | 2,053 | 16,996 | 12 | 2,104 |
| Unique entities | 2,053 | 16,996 | 660 | 372,590 |
| Time periods | 9 quarters | 9 quarters | 3 years | 9 quarters |
| Data frequency | Quarterly | Quarterly | Annual | Quarterly |
| *Change Profile* |  |  |  |  |
| Change prevalence | 1.4% | 24.1% | 74.9% | 14.9% |
| Change events, $n$ | 233 | 36,672 | 2,445 | 440,581 |
| Increases / Decreases | 97.8 / 2.2% | 45.6 / 54.4% | 70.9 / 29.1%<sup>†</sup> | 99.7 / 0.3% |
| Mean $\vert \Delta\vert $ | \$948 | \$2.29 | \$50.27 | \$14.70 |
| Median $\vert \Delta\vert $ | \$385 | \$1.12 | \$20.49 | \$1.97 |
| *Methodology* |  |  |  |  |
| Validation design | 70/15/15<br>Group-aware | 70/15/15<br>Group-aware | LOGO-CV<br>(12 folds) | 70/15/15<br>Group-aware |
| Grouping variable | HCPCS | HCPCS | HCPCS | HCPCS |
| Repeated runs | 30 | 30 | 12 folds | 30 |
| Entity key | HCPCS | HCPCS | HCPCS +<br>Contractor | HCPCS + Modifier<br>+ State + Rural |

<sup>†</sup> AFS overall direction: 70.9% increases / 29.1% decreases. Mileage codes (A0425, A0435, A0436) show 100% rate increases; non-mileage codes show bidirectional changes.
CLFS = Clinical Laboratory Fee Schedule; PFS = Physician Fee Schedule; AFS = Ambulance Fee Schedule; DMEPOS = Durable Medical Equipment, Prosthetics, Orthotics and Supplies; HCPCS = Healthcare Common Procedure Coding System; LOGO-CV = Leave-One-Group-Out Cross-Validation; $|\Delta|$ = absolute magnitude of rate change.
Model-ready rows exclude first-period observations with no lag history. All data downloaded from cms.gov (2024–2026). No patient-level data used.

### 2.2 Validation Framework

To evaluate FeePredict’s generalizability to procedure codes unseen during training, a group-aware approach was used for three of the four fee schedules (CLFS, PFS, and DMEPOS). The HCPCS codes were used as the grouping variable, with all rows with the same procedure code assigned to the training, validation, and test sets, and no group split across those partitions. This strategy avoids situations in which the model may memorize examples with a particular code during training, and then use those memorized parameters to appropriately predict fees for that same code in a later partition of the data. Each dataset was split into 70% training, 15% validation, and 15% test sets by HCPCS grouping. The validation data was used to determine the threshold at Stage 1, and the metrics were calculated on the unseen test data. These splits were performed 30 times with different random seeds to evaluate stability. For visualizations of single-run model output, such as the predicted-versus-actual scatter plots in Figure 3, we display the run whose concordance index equaled the median across the 30 seeds, so that the visualization reflects typical rather than best-case or worst-case performance. Performance metrics reported in Section 3 and in Table 4 aggregate across all 30 runs (mean *±* SD). Additionally, an out-of-time validation was performed on CLFS, PFS, and DMEPOS, which is described in Section 3.2.

A zero-change baseline was calculated for each fee schedule under each evaluation condition. For each row in the dataset, the model predicts no change in the rate for all observations. This indicates the prediction the baseline model would make if it lacked predictive capabilities. All performance metrics are relative to this baseline.

Because the Ambulance Fee Schedule contained only 12 unique HCPCS codes, it was not possible to create a 70/15/15 split of the data. Thus, leave-one-group-out cross-validation was performed with HCPCS codes as the grouping variable, yielding 12 folds. Each fold used one of the procedure code groups as the test set, while the remaining groups were used to train the model. An out-of-time validation was not performed on the Ambulance Fee Schedule due to its annual data collection frequency and three-year observation window.

### 2.3 Leakage Prevention

The data used for this model is time-series data broken down by codes. With time series data, one of the major issues that can arise is what is referred to as leakage, where the model is provided with future information that essentially gives it the answer to the outcome at the present time, inflating the model’s true “power” to appropriately predict the outcome at any given time. To avoid this issue, all features were lagged by one time period (whether that be a year or quarter). Thus, the model would only have knowledge of information from the last quarter and prior information, and would not have access to any information present during the time period or to any future quarter information. This lag information was included in the FeePredict Lag-1 column of the dataset and used for features related to rate changes, changes in rolling statistics, change frequency, the last time the feature changed, and additional features related to the fee schedule, such as the ceiling and floor prices for DMEPOS. To demonstrate that these lag features were in effect, leaky versions of these features were calculated, identical to the lag features but with no lag information included. While the script was running, these two features printed and confirmed that the leaky and lagged information within each feature had almost entirely different rows, indicating that the lag features were active and working.

Additionally, to confirm that there was no leakage across any of the models run, a label-shuffle sanity test was performed for each fee-schedule model. The procedure was as follows: the outcome labels in the training set were randomly permuted, breaking any relationship between features and outcomes, a fresh model was trained on this permuted training set, and that model was evaluated on the unshuffled test set. This shuffle-train-evaluate procedure was repeated 50 times with a different random permutation each time, producing a null distribution of AUC values that represents the performance achievable by a model trained on noise. Any leakage in the model would allow it to learn these randomly permuted outcomes, inflating the null AUC well above 0.5. If there is no leakage, the model has no basis for prediction and the null AUC should remain near 0.5. The mean null AUCs across the 50 shuffles were 0.467 for CLFS, 0.501 for PFS, 0.515 for AFS, and 0.486 for DMEPOS, all near the 0.5 chance baseline, indicating the absence of leakage in the models tested.

### 2.4 FeePredict Pipeline Architecture

The unique four fee-schedule full-predictive-model pipelines can be broken down into three modular models with slight variations to account for the different changes each fee schedule may require. The three models are the Change Classifier, the Direction Classifier, and the Magnitude Regression model. The predicted positives from the Change Classifier will be routed to the Direction and Magnitude Classifier models to determine the change to each fee schedule. Any rows that are classified as negative or no change from the Change Classifier will be assigned a predicted change of zero to the fees.

**Stage 1** classifies whether each fee schedule will change. The model’s inputs are all features deemed safe and lagged from each fee schedule. The classifier’s output is a probability score of the change that will occur to each fee schedule. For the classifier threshold, the optimal F1 point is selected on the validation data set. Furthermore, the model was trained to account for class imbalance within each fee schedule. For instance, the prevalence of change events across fee schedules ranges from 1.4% for CLFS to 74.9% for AFS. A Random Forest Classifier was used for this stage, with hyperparameters varied across models (further explained in Section 2.7).

**Stage 2** determines, for each event with a positive Change Classifier prediction, whether the change in the fee schedule will be an increase or a decrease from the current rate. This model is trained only on the rows that have a change in the fee schedule in the training set. Furthermore, the vast majority of changes in CLFS and DMEPOS were fee increases, leading to a nearly degenerate classification problem for these two fee schedules. Yet, Stage 2 is still included in each fee schedule’s classification pipeline to provide consistency in the models. However, it contributes minimally to these two fee schedules. By contrast, Stage 2 contributes more importantly to PFS and AFS, where true increases and decreases are equally likely. Again, the Random Forest Classifier is used for Stage 2, with hyperparameters varied across models (as further explained in Section 2.7).

**Stage 3** determines the magnitude of change to the fee schedules. Similar to Stage 2, the magnitude regression is trained only on instances where the fees change. Furthermore, the magnitude is incredibly right-skewed between fee schedules. Thus, log1p is applied to the magnitude to normalize the changes, and a Random Forest Regressor is used to estimate the magnitude change. This can be quite the challenge as the mean absolute change in fees across the four fee schedules ranges from $2.29 for PFS to $948 for CLFS. The log1p transform is reverted to the magnitude of the fee change using expm1. Here, a Random Forest Regressor was used for Stage 3, and the hyperparameters varied across fee schedules (as further explained in Section 2.7).

The full model pipeline combines the three stages into one model for each fee schedule. The magnitude of the change will be Stage 2’s classified change direction multiplied by the magnitude estimation of Stage 3’s model for any row with a Stage 1 positive outcome. Furthermore, all rows classified by Stage 1 as ‘no change’ will be assigned a fee change of 0. Prior to training, each stage for each fee schedule was median-imputed and scaled using a StandardScaler. The numbers used to preprocess this data were calculated only from the training rows, and those same numbers were used for transform-only when applying the model to the validation and test data to avoid data leakage.

Random Forest was selected as the underlying model family for all three stages based on three considerations. First, the data is tabular with mixed feature types (continuous rates and RVUs, encoded categorical fields, and binary flags), a setting in which tree ensembles consistently match or outperform alternatives such as support vector machines and neural networks on benchmark studies of tabular data [4, 5]. Second, the temporal structure of the problem is already captured through engineered lag and rolling features rather than through latent state dynamics, making feed-forward tree ensembles a more natural fit than sequence models such as Hidden Markov Models. Third, Random Forest provides native feature importance estimates, which support the interpretability needs of a paper whose conclusions depend partly on identifying which fee schedule features (e.g., the PFS conversion factor, DMEPOS competitive bidding) drive predictability. Formal benchmarking against alternative model families was outside the scope of this study and is identified as a direction for future work in Section 5.

The pipeline architecture is illustrated in Figure 1.

**Figure 1:**
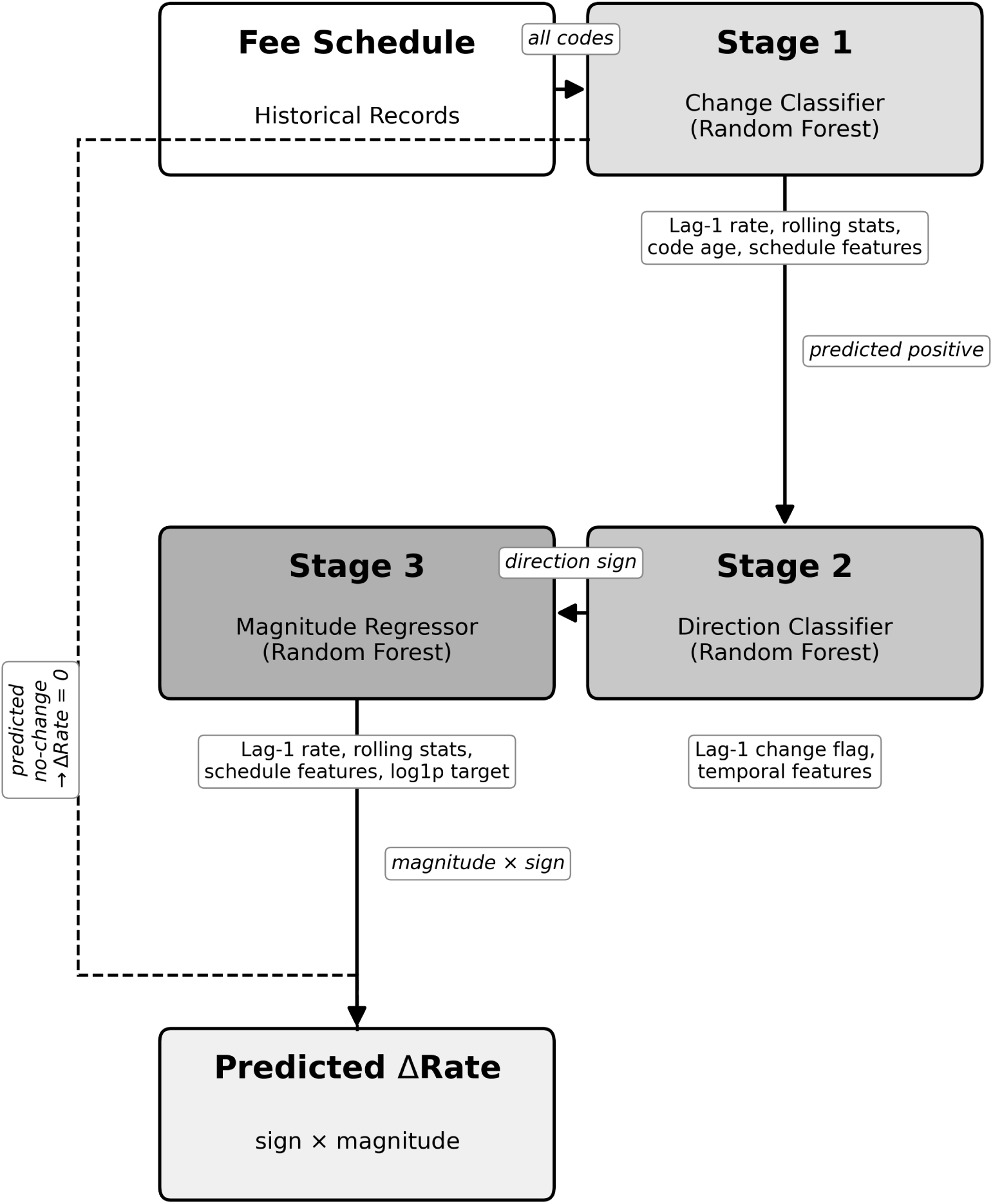
FeePredict three-stage pipeline architecture. Predicted positives from Stage 1 are routed to Stages 2 and 3; predicted no-change rows are assigned a delta of zero. The pipeline is applied independently to each of the four Medicare fee schedules.

**Figure 2:**
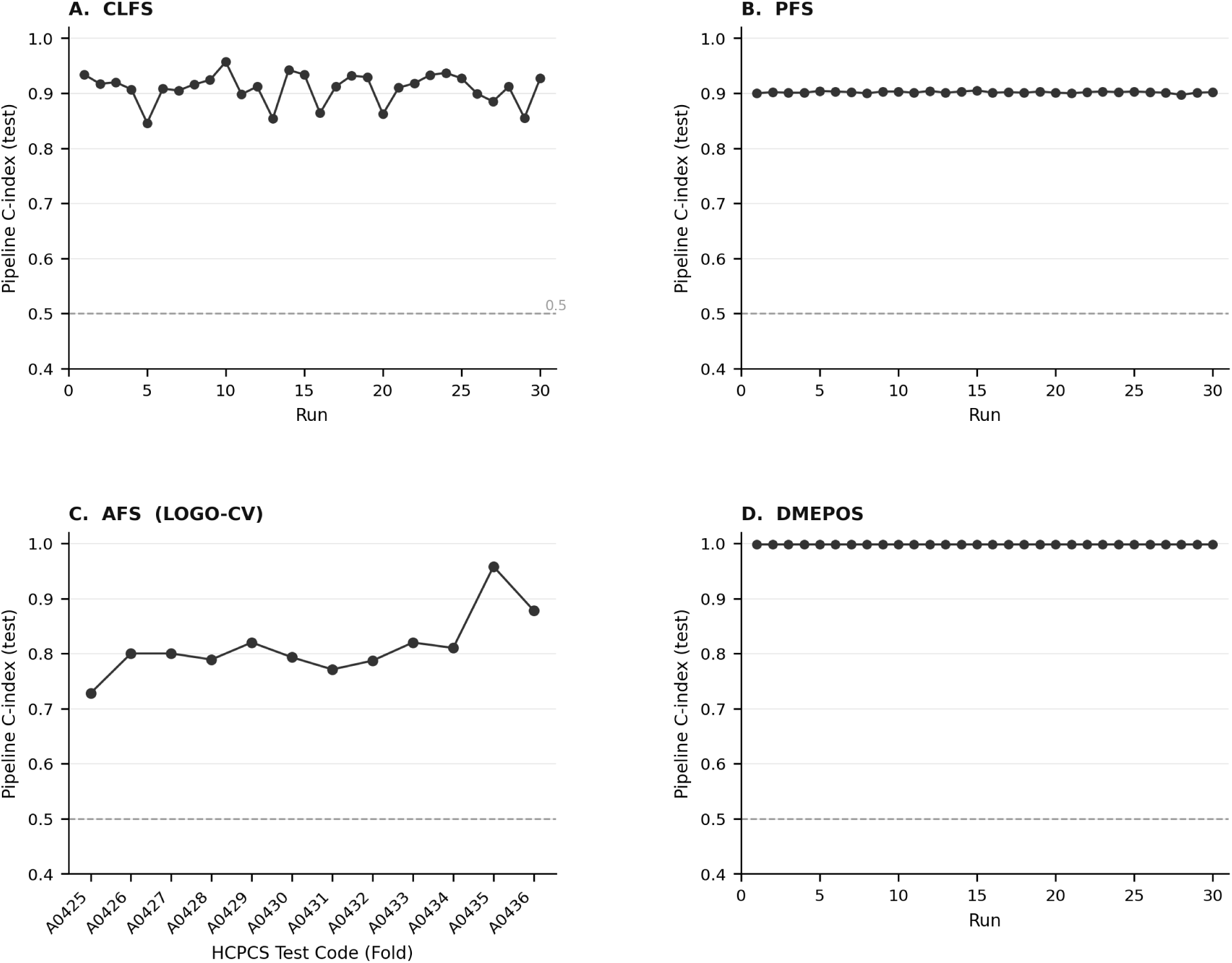
Pipeline concordance index (C-index) across runs for each fee schedule. (A) CLFS: 30 random splits. (B) PFS: 30 random splits. (C) AFS: LOGO-CV by HCPCS test code (12 folds). (D) DMEPOS: 30 random splits. Dashed line indicates chance performance (C-index = 0.5).

**Figure 3:**
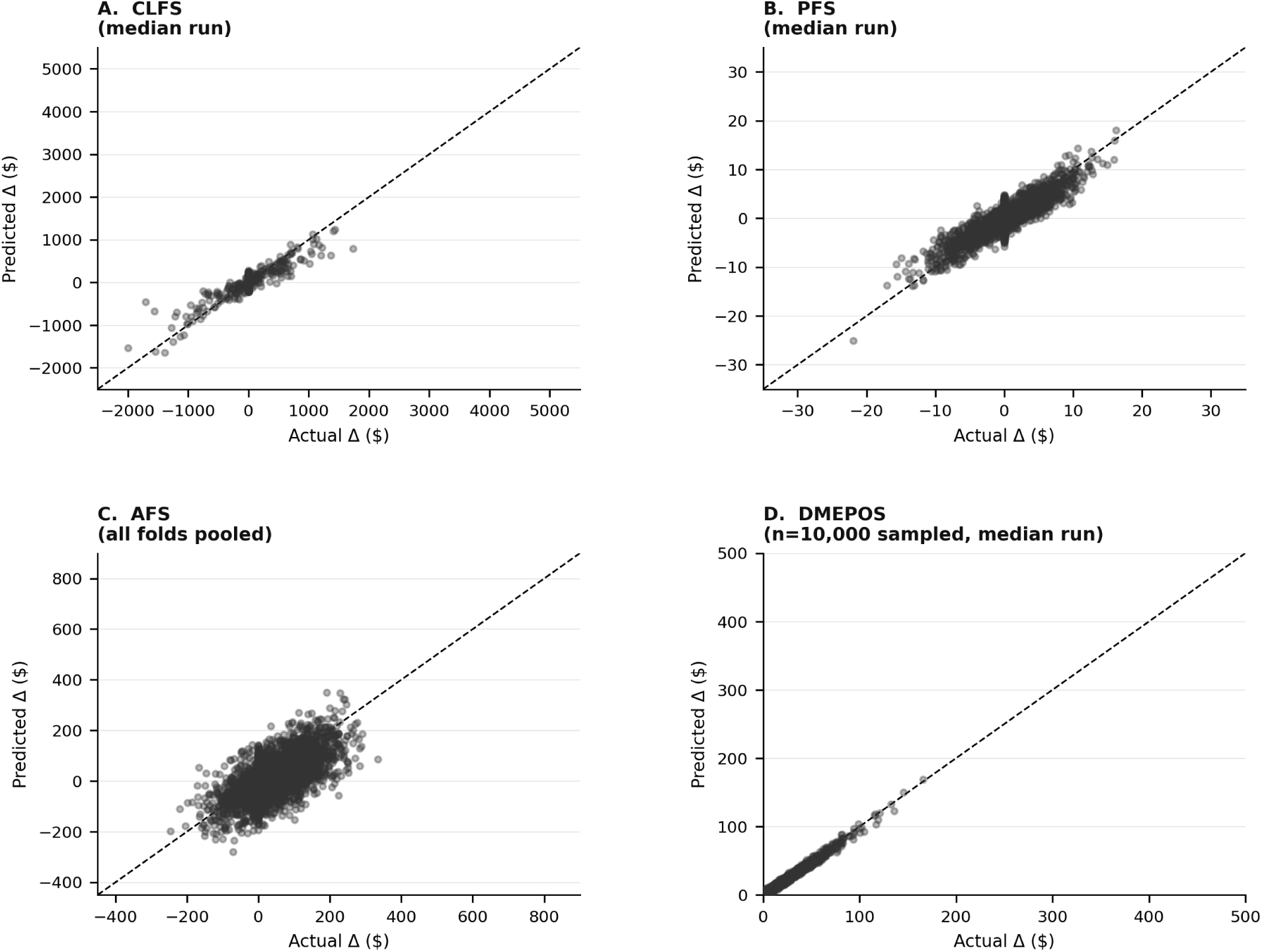
Pipeline predicted versus actual rate change (Δ) for the representative run across all four fee schedules. (A) CLFS (Run 29). (B) PFS (Run 22). (C) AFS (all folds pooled). (D) DMEPOS (Run 30; *n* = 10,000 sampled). Dashed line indicates perfect prediction.

**Figure 4:**
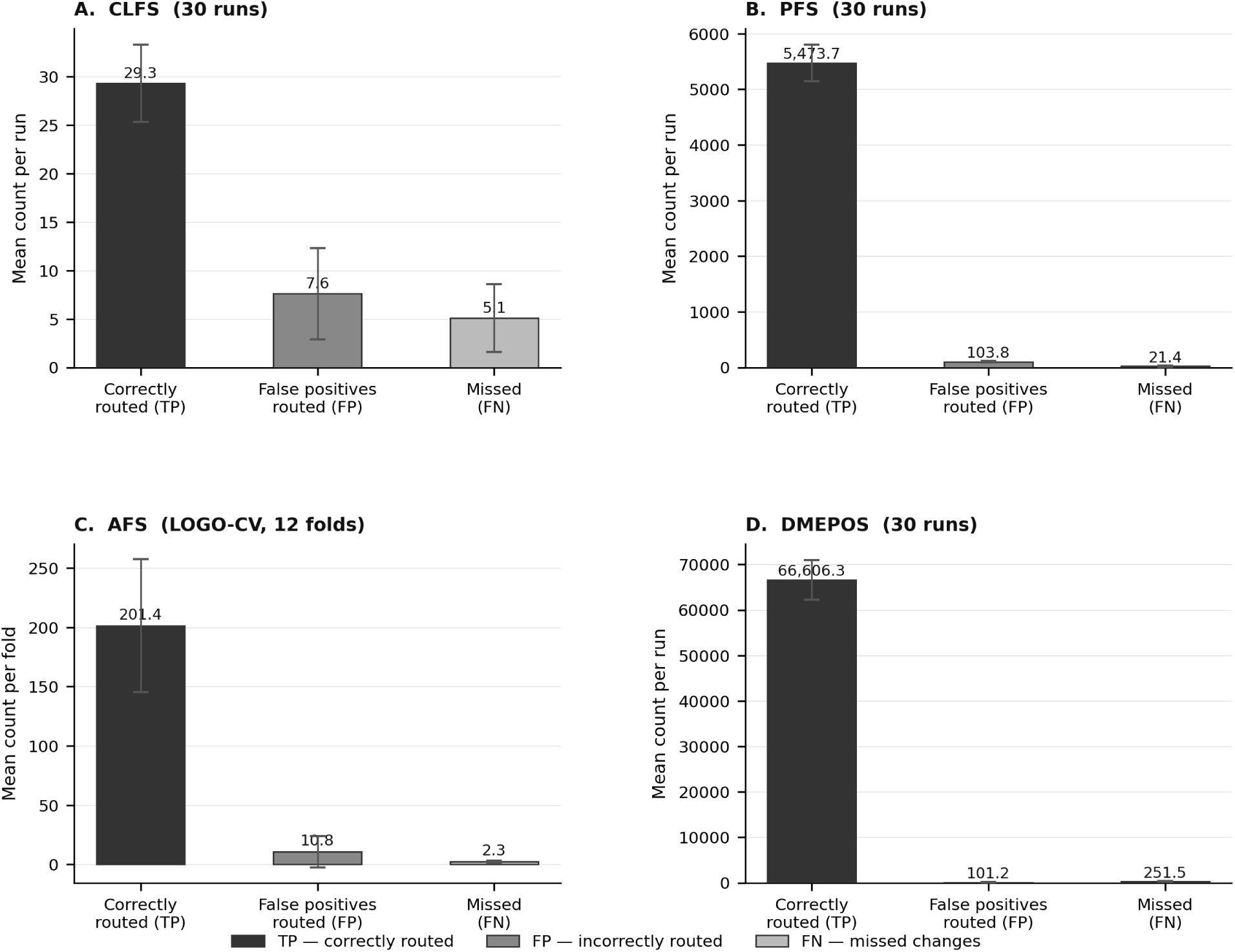
Stage 1 error propagation analysis across all four fee schedules. Bars show mean counts of correctly routed true positives (TP), false positives routed to Stages 2 and 3 (FP), and missed true changes (FN). Error bars represent *±*1 SD. (A) CLFS: mean *±* SD across 30 runs. (B) PFS: mean *±* SD across 30 runs. (C) AFS: mean *±* SD across 12 LOGO-CV folds. (D) DMEPOS: mean *±* SD across 30 runs.

### 2.5 Schedule-Specific Features and Design Decisions

All fee schedules utilize the same universal base features computed from each fee schedule’s dataset, as well as some features specific to that fee schedule model. The 12 base features that each model includes, with the exception of the AFS feature, include features that represent information about the rate itself, such as the year, quarter, code age in quarters and years, the rate lag1 (which is the lag1 of the urban rate for the AFS), lag-1 change history (which is represented by the changed flag for lag1 as well as the quarters and years since the rate had changed), and various rolling statistics for the rate (such as the rolling mean, standard deviation, and change frequency for rates calculated over four and eight quarters periods while the AFS utilizes 2-year rolling periods instead). Other features are created for each fee schedule based on its strengths, as they have inherent predictive power for those specific features. For instance, the CLFS includes 6 additional text-derived features, bringing its total to 18. The PFS uses a conversion factor lag, RVU, and code structural features to account for its annual conversion factor update, bringing its total features to 27. For the AFS, features such as GPCI locality, rate structure, service type, and geographical features are created to account for adjustments by locality and service type, bringing the total to 23 features. For the DMEPOS codes, features such as the equipment modifier, competitive bidding, and structural features are created to account for competitive bidding and equipment type, for a total of 24 features. Further information regarding these features is presented in Table 2.

**Table 2:** Feature categories across all four Medicare fee schedules. Values represent the number of features in each category per schedule. The full feature list with individual feature names and Stage 2 exclusions is provided in Supplementary Table S1.

| Feature Category | CLFS | PFS | AFS | DMEPOS |
| --- | --- | --- | --- | --- |
| *Base Features (all schedules)* |  |  |  |  |
| Temporal | 3 | 3 | 2 | 3 |
| Lag-1 rate & change history | 2 | 2 | 3 | 2 |
| Rolling statistics | 6 | 6 | 4 | 6 |
| *CLFS-Specific* |  |  |  |  |
| Text-NLP (description) | 5 | — | — | — |
| Code structural | 1 | — | — | — |
| *PFS-Specific* |  |  |  |  |
| Conversion factor (CF) | — | 3 | — | — |
| RVU features | — | 4 | — | — |
| Code structural | — | 5 | — | — |
| Text-NLP (description) | — | 2 | — | — |
| *AFS-Specific* |  |  |  |  |
| GPCI locality adjustment | — | — | 3 | — |
| Rate structure (urban/rural) | — | — | 3 | — |
| Service type | — | — | 4 | — |
| Geography (locality) | — | — | 2 | — |
| Rolling statistics (rural) | — | — | 2 | — |
| RVU | — | — | 1 | — |
| *DMEPOS-Specific* |  |  |  |  |
| Equipment modifier | — | — | — | 4 |
| Competitive bidding | — | — | — | 4 |
| Structural / geographic | — | — | — | 4 |
| **Total features** | **18** | **27** | **23** | **24** |

Values represent the number of features in each category per schedule. — = category not applicable to this schedule.
Full feature list with individual feature names and Stage 2 exclusions provided in Supplementary Table S1.
\* AFS base features use annual variants: 2-year rolling windows, `urban_rate_lag1`, `years_since_last_change`. QUARTER not included.

The entity key for the data helps determine which time series is considered unique. If the entity key had been defined incorrectly for each schedule, the lag-1 operation could have crossed boundaries that were not supposed to be crossed for that schedule, thereby fabricating changes in the rates that did not actually occur. Only two fee schedules required specific entity key decisions: the AFS and DMEPOS schedules. For the AFS, the entity key was defined as the HCPCS code as well as the contractor and carrier for that code, but only for codes that did not have localities specifically associated with them – there were 18 contractors that had multiple localities, thus excluding those based on this feature would prevent potential issues with the model. For the DMEPOS codes, the entity key included the HCPCS code, modifier 2, the state in which the codes are used, and whether the codes were for a rural or urban area. This specific feature was included because a modifier could cross the same HCPCS code, leading to different prices for the same code (e.g., renting versus purchasing the same equipment).

Additionally, both the PFS and DMEPOS fee schedules have a Stage 1 AUC of 1.000. These are not due to label leakage but due to the structure of the fee schedules. For the PFS, the conversion factor changes every January, affecting approximately 49% of all codes in that fee schedule in the Q1 quarter alone, compared to only 14.5% in another quarter of the year. For the DMEPOS codes, the rural codes with a zero reimbursement rate (which represent 50% of all DMEPOS codes) do not change at all, while the non-rural codes experience the same update as the PFS. Thus, for both the PFS and DMEPOS codes, the model identified these structural features. The label-shuffle test with null AUCs of 0.515 for PFS and 0.486 for DMEPOS also provided evidence of the absence of leakage in the given experiments.

### 2.6 Evaluation Metrics and Statistical Tests

#### Individual Stage Metrics

In Stage 1, the AUC and F1 score are utilized. The AUC can help measure discrimination between the positive and negative classes, even with class imbalance. The F1 score provides insight into how the model performs at the chosen threshold, whereas the AUC is threshold-independent. In Stage 2, the AUC is used to assess the model’s ability to rank instances where the target variable increases versus those where it decreases. This metric is essentially meaningless for CLFS and DMEPOS, as the relationship between the target and features for those fee schedules is almost unidirectional. In Stage 3, three different metrics are used: mean absolute error (MAE), Spearman correlation, and *R*^2^. The MAE was chosen over the RMSE because the distribution of target values is right-skewed. The MAE is less sensitive to outliers than the RMSE. Furthermore, the Spearman correlation is also robust to outliers and can help measure the ranking ability of the model’s estimated magnitudes for each code. Finally, the *R*^2^ statistic is reported for completeness of the model evaluation, but is sensitive to outliers, which should be noted given the distribution of the target variable magnitudes.

#### Full Pipeline Metrics

The evaluation metrics for the end-to-end pipelines for each fee schedule are the Concordance Index (C-Index), MAE, and the Spearman correlation statistic applied to the change rows. The C-Index indicates the model’s ability to rank a randomly selected record in the positive class (codes that change) higher than a randomly selected record in the negative class (codes that do not change). The C-Index was calculated using Kendall tau to improve computational efficiency compared to calculating the concordance index via the ROC AUC. The mean absolute error was calculated for all rows and again only for the rows that indicate a change in codes. The Spearman correlation coefficient was calculated again for the same reasons as in Stage 3.

#### Statistical Tests

Two statistical tests were performed to evaluate the robustness of the models. The first test was a one-sided Z-test against the null hypothesis that the pipeline Concordance Index equals 0.5 (chance performance). For each fee schedule, the test statistic was calculated as

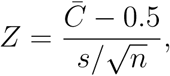

where *C̅* is the mean pipeline C-index across the *n* = 30 cross-validation runs (12 folds for AFS), *s* is the standard deviation of the C-index across those runs, and 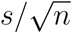 is the standard error of the mean. Because the runs share underlying data and are not strictly independent, this test should be interpreted as a coarse robustness check rather than a formal hypothesis test. The label-shuffle test described in Section 2.3 provides a complementary, assumption-free null comparison. The second test was the label-shuffle test for sanity, which was described in Section 2.3.

### 2.7 Implementation Details

Python version 3.13.3 was used for all modeling and data processing. scikit-learn was used for the three random forest models, along with GroupShuffleSplit, StandardScaler, and median imputation for preprocessing the data. Pandas and NumPy were vital components of the project, serving as the backbone of data manipulation. Kendall’s tau from SciPy was used to calculate the C-Index for the fee schedule models, due to its *O*(*n* log *n*) runtime compared to the naive pairwise-comparison implementation (*O*(*n*^2^)). This was very useful for evaluating the DMEPOS fee schedule, which contained millions of rows of data. Lastly, Matplotlib was used to plot figures for each model.

All experiments were performed on an Apple MacBook Pro M4 with 8 GB of memory. The runtimes for each fee schedule model differed due to differences in dataset size. The AFS was the fastest, taking only a few minutes to complete. Following the CLFS model, which took only slightly longer to calculate, the PFS took only a minute or two longer. The longest runtime was for the DMEPOS model, which took approximately 60 minutes to complete all 30 analysis runs.

To ensure model reproducibility, two steps were taken. First, the `PYTHONHASHSEED` was set to 42 for all runs. Additionally, the random split parameter was seeded with RANDOM SEED + run id *×* 17 to ensure that each model (and each run) could reproduce the metrics calculated during the original implementation of the models.

Each raw data file for each year was first downloaded from the website https://www.cms.gov. These individual tables were merged into four different data tables, one for each of the four fee schedule models. Each of these tables was passed through a feature engineering script, producing four CSV files containing the features for each fee schedule model. Each of these feature CSV files was passed through the scripts for each model.

The hyperparameters for each random forest model and fee schedule are reported in Table 3. These hyperparameters were set empirically to account for the dataset size. A formal search for hyperparameters was not performed, but this is an area for future optimization of these models’ performance.

**Table 3:** Random forest hyperparameters for all three pipeline stages across all four fee schedules. Parameters were set empirically based on dataset scale and computational constraints.

| Parameter | CLFS |  |  | PFS |  |  | AFS |  |  | DMEPOS |  |  |
| --- | --- | --- | --- | --- | --- | --- | --- | --- | --- | --- | --- | --- |
|  | S1 | S2 | S3 | S1 | S2 | S3 | S1 | S2 | S3 | S1 | S2 | S3 |
| `n_estimators` | 800 | 500 | 300 | 500 | 300 | 300 | 500 | 300 | 300 | 200 | 200 | 200 |
| `max_depth` | None | 5 | 6 | 10 | 8 | 8 | 6 | 5 | 5 | 10 | 8 | 10 |
| `min_samples_split` | 2 | 4 | 4 | 4 | 4 | 4 | 4 | 4 | 4 | 10 | 10 | 10 |
| `min_samples_leaf` | 1 | 2 | 3 | 2 | 2 | 3 | 2 | 2 | 3 | 5 | 5 | 5 |
| `max_features` | sqrt | sqrt | sqrt | sqrt | sqrt | sqrt | sqrt | sqrt | sqrt | sqrt | sqrt | sqrt |
| `class_weight` | bal. | bal. | — | bal. | bal. | — | bal. | bal. | — | bal. | bal. | — |
| `bootstrap` | True | True | True | True | True | True | True | True | True | True | True | True |
| Model type | RFC | RFC | RFR | RFC | RFC | RFR | RFC | RFC | RFR | RFC | RFC | RFR |

S1 = Stage 1 Change Classifier; S2 = Stage 2 Direction Classifier; S3 = Stage 3 Magnitude Regressor. RFC = RandomForestClassifier; RFR = RandomForestRegressor.
bal. = `class_weight='balanced'` (S1 and S2 only); — = not applicable (regression); None = unconstrained tree depth.
All models: `max_features='sqrt'`, `bootstrap=True`, `n_jobs=-1`. Parameters were set empirically based on dataset scale and computational constraints. A formal hyperparameter search was not conducted.

## 3 Results

### 3.1 Cross-Schedule Pipeline Performance

The FeePredict analysis shows that the model outperforms the zero-change baseline across all four fee schedule structures and for all four training and testing schedules. For clarity, the metrics reported include the area under the curve (AUC) for Stages 1 and 2, which measures model discrimination between fee changes and non-changes; a value of 1 indicates perfect discrimination, and 0.5 indicates random guessing. Stage 1 AUC values range from 0.975 to 1.000, and Stage 2 AUC values range from 0.614 to 0.969. The concordance index (C-index), a metric for predictive accuracy in ranking outcomes, ranges from 0.815 (AFS) to 0.998 (DMEPOS). Mean absolute error (MAE), which measures the average error between predicted and observed fee changes, shows improvements relative to the baseline, ranging from 18% for the CLFS fee schedule to 85% for the DMEPOS fee schedule. For only rows with a change in the fee schedule, the MAE improvement ranged from 29% for CLFS to 85% for DMEPOS.

AUC was calculated for each of the four fee schedules for each of the two stages of the models. For instance, for the CLFS fee schedule for Stage 1, the AUC was 0.975*±*0.019, with a label-shuffle null AUC of 0.467; for Stage 2, the AUC was 0.614. For the PFS fee schedule, the AUC for Stage 1 was 1.000 *±* 0.000, with a null AUC of 0.501 for Stage 2; the AUC was 0.956 *±* 0.004. For the AFS fee schedule, the AUC for Stage 1 was 0.994 *±* 0.003, with a null AUC of 0.515 for Stage 2; the AUC for Stage 2 was 0.969 *±* 0.006. For the DMEPOS fee schedule for Stage 1, the AUC was 0.999 *±* 0.001, with a null AUC of 0.486. For Stage 2, the AUC was 0.850 *±* 0.182.

Each fee schedule also included individual C-indices for each model. For instance, the CLFS fee schedule yielded a C-index of 0.912 *±* 0.030 and a Z-test p-value of *<* 0.001. The PFS fee schedule returned a C-index of 0.903 *±* 0.006, with a Z-test p-value of *<* 0.001. The AFS fee schedule returned a C-index of 0.815 *±* 0.057, with a Z-test p-value *<* 0.001. Lastly, the DMEPOS fee schedule yielded a C-index of 0.998 *±* 0.001 and a Z-test p-value of *<* 0.001. The C-index values for each fee schedule were all well above 0.5, the chance that a model with two procedure codes will correctly predict which procedure has a larger rate of change in the next period. Thus, the model works well across all four fee schedule structures.

MAE improvements varied across fee schedules (Table 4). For the CLFS, the pipeline achieved an MAE of $10.84 across all rows and $683 for change rows, compared to baselines of $13.19 and $959, respectively. This corresponds to an 18% improvement in MAE across all rows and a 29% improvement for change rows. For the PFS, the pipeline MAE was $0.22 across all rows and $0.89 for change rows, compared to baselines of $0.55 and $2.29, yielding improvements of 60% and 61%, respectively. For the AFS, the pipeline MAE was $23.76 across all rows and $26.45 for change rows, compared to baselines of $32.69 and $43.63, corresponding to improvements of 27% and 39%. For the DMEPOS, the pipeline MAE was $0.34 across all rows and $2.16 for change rows, compared to baselines of $2.20 and $14.70, yielding an 85% improvement in both cases.

**Table 4:** Cross-schedule pipeline performance summary. All metrics reported as mean *±* SD across 30 runs (LOGO-CV for AFS: 12 folds). Improvement denotes reduction in MAE relative to the zero-change baseline. All Z-tests: one-sided test of pipeline C-index vs. random (0.5), *p <* 0.001.

| Metric | CLFS<br>(n=16,638) | PFS<br>(n=152,202) | AFS<br>(n=3,264) | DMEPOS<br>(n=2,952,842) |
| --- | --- | --- | --- | --- |
| *Stage 1 — Change Detection* |  |  |  |  |
| AUC (mean ± SD) | 0.975 ± 0.019 | 1.000 ± 0.000<sup>†</sup> | 0.994 ± 0.003 | 0.9999 ± 0.0001<sup>†</sup> |
| Label shuffle null AUC | 0.467 | 0.501 | 0.515 | 0.486 |
| *Stage 2 — Direction Classification* |  |  |  |  |
| Direction AUC (mean ± SD) | 0.614<sup>‡</sup> | 0.956 ± 0.004 | 0.969 ± 0.006<sup>§</sup> | 0.850 ± 0.182 |
| *Pipeline — End-to-End Performance* |  |  |  |  |
| C-index (mean ± SD) | 0.912 ± 0.030 | 0.903 ± 0.006 | 0.815 ± 0.057 | 0.998 ± 0.001 |
| Z-test vs random (p) | <0.001 | <0.001 | <0.001 | <0.001 |
| *MAE — All Rows* |  |  |  |  |
| Pipeline MAE (\$) | \$10.84 | \$0.218 | 23.76/12.64<sup>¶</sup> | \$0.340 |
| Baseline MAE (\$) | \$13.19 | \$2.282 | — | \$2.199 |
| **Improvement** | **18%** | **90%** | — | **85%** |
| *MAE — Change Rows Only* |  |  |  |  |
| Pipeline MAE (\$) | \$683 | \$0.893 | \$26.45 | \$2.157 |
| Baseline MAE (\$) | \$959 | \$2.282 | \$43.63 | \$14.700 |
| **Improvement** | **29%** | **61%** | **39%** | **85%** |
| *Dataset Characteristics* |  |  |  |  |
| Change prevalence | 1.4% | 24.1% | 74.9% | 14.9% |
| Direction (inc. / dec.) | 97.8 / 2.2% | 45.6 / 54.4% | 70.9 / 29.1% | 78.3 / 21.7% |
| Evaluation design | 30-run CV | 30-run CV | LOGO-CV (12) | 30-run CV |

CV = cross-validation with group-aware 70/15/15 splits; LOGO-CV = leave-one-HCPCS-out cross-validation. <sup>†</sup> Stage 1 AUC reflects structural near-perfect separability confirmed by label shuffle (null AUC ≈ 0.50); not data leakage. <sup>‡</sup> CLFS Stage 2 AUC reported at pipeline level; activation/deactivation events structurally linked to direction (is\_activation excluded from Stage 2 features). <sup>§</sup> AFS Stage 2 AUC for non-mileage folds only; mileage codes (A0425, A0435, A0436) show 100% directional increases (direction trivial). <sup>||</sup> DMEPOS Stage 2 AUC shows high run-to-run variance ( ± 0.182) attributable to modifier class imbalance in random splits; median across runs = 0.952. <sup>¶</sup> AFS all-row MAE: mean / median. High variance driven by air transport codes (A0430, A0431); median is more representative. Improvement = reduction in MAE relative to zero-change baseline. All Z-tests: one-sided test of pipeline C-index vs random (0.5), $p < 0.001$ .

Across all four schedules, the change-row MAE is the more operationally meaningful metric, because it isolates predictive performance on the subset of codes for which rates actually move and removes the dilution introduced by the large fraction of unchanged codes. Subsequent analyses therefore focus on the change-row MAE as the primary measure of pipeline accuracy.

The predictability of each of these fee schedule structures varied due to their different structures. For instance, the DMEPOS fee schedule showed the greatest improvement in MAE for only rows that changed fees (85%), due to the use of competitive bidding, a feature present only in the DMEPOS fee schedule. The PFS fee schedule showed an improvement in MAE of 61% for only the rows that changed fees, due to the use of a conversion factor published and announced each year for all procedures within the PFS fee schedule. The AFS fee schedule showed an improvement in MAE of 39% for only the rows that changed fees, because it uses a GPCI value that can vary from locality to locality within the fee schedule. Finally, the CLFS fee schedule showed the lowest improvement in MAE for only rows that changed fees (29%), because the rates for clinical laboratory tests are set to a non-zero rate rather than 0, a change that is difficult to learn, yet is reflected in model predictions for CLFS tests.

### 3.2 Out-of-Time Generalization

An out-of-time (OOT) validation was used for the CLFS, PFS, and DMEPOS models, where each model was trained on all available years in the past and then evaluated on a held-out period in the future. AFS was not included in this evaluation due to its different evaluation methodology (LOGO-CV) and limited time windows for its data (3 total). The OOT validation is the most relevant form of generalization for models, as it evaluates their ability to accurately predict outcomes in a forward period.

CLFS and DMEPOS both achieved strong results during their OOT validation periods, indicating strong time generalization. For CLFS, the C-Index dropped to 0.854 during the OOT period, compared with its original C-Index of 0.912 for cross-validation testing. While this is a small drop in performance (0.058), it is to be expected given the difficulty of predicting CPT code activation in the near future. DMEPOS experienced an even smaller drop in its C-Index, from 0.998 in the cross-validated test to 0.972 during OOT validation, indicating the stability of the adjustments to DMEPOS’s competitive bidding rates.

PFS, the only fee schedule model to show a substantial drop in performance during the OOT validation period, dropped to a C-Index of 0.592 from its original C-Index of 0.903 during cross-validation testing. While the AUC for PFS remained the same during out-of-time validation (AUC = 0.974), the AUC for Stage 2 of PFS dropped from 0.956 to 0.703. The reason for this drop most likely aligns with the PFS conversion factor update during the OOT validation period. Rates for PFS are calculated by multiplying RVUs by a conversion factor; because the conversion factor was not the standard conversion factor, the PFS model was unable to accurately predict the magnitude of RVUs for the forward and unseen periods. This type of change is referred to as a regime shift, which cannot be detected during cross-validation. These results motivate the recommendation that the PFS model be recalibrated each year after CMS announces the new conversion factor, which is published months before the factor’s effective date (Further discussion in Section 4).

### 3.3 Error Propagation Analysis

Every FeePredict fee schedule model operates through a sequential three-stage pipeline, in which errors in Stage 1 can propagate to affect outcomes in Stages 2 and 3 as well. For example, false negatives in Stage 1 would result in the predicted rate change delta for all affected codes being 0, regardless of the actual rate change in those codes in Stage 3. Furthermore, false positives in Stage 1 result in the codes that do not experience any rate change being passed through to Stage 2 and 3 in the pipeline, again resulting in a large difference between the actual rate change of 0 and the predicted rate change delta. Each of these error rates can be quantified by tracking counts of true positives, false positives, and false negatives.

For the PFS, AFS, and DMEPOS fee schedules, the counts of routing errors in Stage 1 are small. For each of these three fee schedules, the recall rate for correctly routed codes to Stage 2 is 98.9% or higher. Thus, only 1.1% or less of the true rate changes to codes are missed by these three fee schedule models. Furthermore, the number of codes correctly classified as having no rate change but routed through to Stages 2 and 3 is small: 1.9% for PFS, 5.1% for AFS, and 0.2% for DMEPOS codes. These small numbers indicate that errors in the fee schedule models are due to the magnitude of the rate changes rather than the routing of codes through the model to Stages 2 and 3. Thus, these errors are appropriate for correcting the fee schedule models in their future implementations rather than for detection within the current model.

However, the counts for these three fee schedules differ significantly from those for the CLFS fee schedule. Its higher routing error rate for codes with no rate change reflects the structural change in CLFS codes that experience rare rate-change events (occurring at a 1.4% prevalence in the dataset). Recall for correctly routed codes that reach Stage 2 of the pipeline is 85.2%, meaning that 34 codes change rate within the test set of 2,400 codes. Thus, on average, the CLFS fee schedule model misses 5.1 true rate changes and incorrectly routes 7.6 codes that do not change their rates to Stage 2 of the model. Despite these errors, however, the performance is still operationally acceptable; it correctly detects 6 out of the 7 true rate changes for codes within the CLFS fee schedule. Thus, the model’s detection errors suggest that its prediction errors for CLFS codes are due to the complexity of the activation events that must occur within the codes, compared with those in the counterpart fee schedules.

## 4 Discussion

### 4.1 Why It Worked and Why It Varied Across Schedules

FeePredict has shown that Medicare reimbursement rates are learnable by a machine learning model using historical data from their current fee schedule; the model’s performance varied across fee schedules, reflecting the individualistic but deterministic manner in which the rates are set. The best performance across the models was with the DMEPOS code entries, achieving a C-index of 0.998 and an 85% reduction in MAE compared to historical reimbursement rates. This was due to the fact that the DMEPOS codes are the only codes within the Medicare system that use competitive bidding, allowing the model to learn the proportional adjustments to the reimbursement rates for each DMEPOS code entry based upon the reimbursement rate for the same code entry during the preceding calendar year (Spearman’s *r* = 0.995). The second-highest-performing fee schedule was the PFS codes, with a C-index of 0.903 and a 61% improvement in MAE. For the PFS codes, reimbursement for each procedure is determined by an annual conversion factor applied to all procedure codes; hence, the model included 3 features representing the lagged values of this conversion factor. The third-highest-performing code was the AFS code, with a C-index of 0.815 and a 39% improvement in MAE. For the AFS codes, the reimbursement is locally adjusted by a GPCI factor that varies from laboratory to laboratory. The AFS local rates (C-Index = 0.822) were more easily predictable than the national average GPCI factor for the AFS codes (C-index = 0.592). The CLFS code entries demonstrated the lowest level of improvement in the C-index (29%) and in the MAE; the reason for this is in the fact that most price changes for the CLFS codes were due to the activation of the code entries, which changed from inactive or a 0 pricing to a priced status for the clinical laboratory services (CLFS) codes. Thus, these changes were not reflected in the historical data provided to the machine learning model.

In addition to examining the performance of the models during training, another important validation study was that of the model’s performance out-of-time (OOT). For the PFS codes, where the model’s cross-validated C-index was 0.903, OOT validation yielded a dramatic drop to 0.592 for the same fee schedule. The Stage 1 AUC scores remained unchanged in the model’s out-of-time validation, but the Stage 2 AUC scores significantly decreased. These results suggest that the model’s ability to rank the magnitude of reimbursement amounts for the procedures within the PFS codes failed during the OOT period. Such a failure may have resulted from an abnormal adjustment to the conversion factor for the PFS codes during the OOT period; in other words, the changes to the reimbursement factor were a regime shift relative to the factor during the model’s training period. Specifically, the CY 2026 PFS final rule (CMS-1832-F, published October 31, 2025) introduced two structural changes absent from the training period: the bifurcation of the single conversion factor into separate QP ($33.57) and non-QP ($33.40) factors, and a *−*2.5% efficiency adjustment applied non-uniformly to the work RVUs of non-time-based services. Because lag-1 feature construction restricted the model’s view to the Q4 2025 conversion factor of $32.35, neither change was visible at prediction time, which is consistent with the observed pattern of preserved Stage 1 discrimination (AUC = 0.974) but degraded Stage 2 magnitude ranking (AUC dropping from 0.956 to 0.703). Another way of stating this finding is that there is a critical characteristic of the policies that govern the reimbursement rates for Medicare code entries and services that is represented by the model results; namely, the reimbursement model is inherently sensitive to changes in policy. This is in contrast to the CLFS and DMEPOS models, which exhibited robustness in their OOT C-index scores relative to their cross-validated training performance, indicating stronger temporal generalization due to their private-payor-rate-anchored laboratory pricing and competitive bidding structures, respectively.

### 4.2 Comparison to Prior Work

The deterministic patterns to which we referred earlier for some of the fee schedules above exist well before our structured framework for understanding them. For instance, Medicare physician reimbursements use a structure for assigning prices to its procedural codes that includes relative value units (RVUs) and a geography-based cost index multiplier. Such a framework, the RBRVS model, was originally devised by Hsiao et al. [1], who divided the work of a physician into intraservice, preservice, and postservice components and developed methods to measure the cost of each component of the physician’s practice. Subsequently, Seidenwurm and Burleson [2] discovered that the conversion factors used by Medicare are updated each year via a formula influenced by the stature of the United States and other macroeconomic factors affecting the country and its population. These adjustments to the conversion factors are the reason for rate changes across all PFS procedural fee schedule codes and are the primary component of the PFS-specific model pipeline.

Exploring the field of machine learning and its applications in healthcare discovery and prediction, we find that it is widely applied in healthcare. However, most of its applications relate to patients within that healthcare facility rather than to understanding the financial systems of healthcare in general, such as Medicare fee schedules. For instance, Morid et al. [3] published a review of the methods for learning about patient costs within healthcare facilities, and found through empirical testing that gradient boosting methods outperformed other machine learning methods in predicting the costs of patients within healthcare facilities, using a data set of approximately 90,000 patients and millions of data points on those patients’ medical and pharmacy claims. Further work has extended this to subpopulation-specific spending adjustment [17] and longitudinal trajectory modeling among initially low-cost patients [19], yet neither addresses reimbursement-rate forecasting at the code level. As such, the approach to machine learning and the predictive power of machine learning models for fee schedule predictions are entirely different problems. While the studies with patients and expenditures focused on the individual predictive nature of patients’ fees and costs, the FeePredict models focus instead on the policy-driven determination of rates for different codes within the Medicare fee schedules. Thus, to our knowledge, FeePredict is the first ML model developed specifically to forecast changes in Medicare fee-schedule reimbursement rates across a variety of fee schedules.

### 4.3 Practical Implications

FeePredict has the potential to be used not just by health systems but also by organizations such as laboratories, ambulance services, and suppliers of durable medical equipment – all of which rely on Medicare revenue to operate. Currently, each of these organizations must manually review the proposed CMS fee schedule changes to produce estimates of the changes to those fee schedules. These manual estimates can potentially lead to errors in revenue projections for those organizations. FeePredict can provide estimates of changes to each procedural code within those fee schedules, allowing organizations to better plan their revenues.

The finding regarding the temporal generalization of the PFS model following the out-of-time validation studies indicates that recalibrating the model after the release of the proposed rule changes to the proposed fee schedule is likely to restore the model’s magnitude ranking performance. The proposed rule changes are typically published in July each year, which coincides with the current review periods for the fees each organization incurs; the only change required at this stage would be to recalibrate stages 2 and 3 of the model.

For the CLFS stage, while the model’s recall is high at 85.2%, there is potential for missed changes to codes, especially within health systems that derive significant revenue from those codes. However, the changes to codes near the end of their three-year private payor rate cycle could be manually reviewed prior to implementation, as they are most likely to change. Regarding the AFS fees, the study results have two implications. First, local fees should replace the national rate for ambulance service providers, as local rates are more accurate. Second, the majority of codes are likely to experience changes in the AFS rates each year. For the DMEPOS codes, the model’s near-perfect temporal generalization indicates that its use does not require recalibration each year for budgeting purposes.

## 5 Limitations

When considering the experiments conducted, a limitation of the data downloaded from the CMS was that it was limited to Medicare fee-for-service data for three years (2024– 2026) across all fee schedules. The results produced in these experiments, therefore, will not necessarily generalize to other Medicaid, commercial payers, or Medicare Advantage plans, each of which has its own specific rate structure. Furthermore, given that the data exists for only a three year period, which does not encompass the full rate cycle for fee schedules such as CLFS that have a three-year rate cycle for private payor repricing, those results will not necessarily be generalizable to future rate cycles due to the potential that the model was not presented with a complete cycle within the data that was presented for model training. Additionally, external validation data were unavailable for these experiments because the CMS was the only data source required to run them. Therefore, this noted limitation in data availability is inherent to the experiments.

Lastly, another limitation of the experiment was the variation in the data provided for the AFS compared to the more symmetrical data for the CLFS, PFS, and DMEPOS fee schedules. Data for the AFS were less temporal in-depth than those for the other three fee schedule codes, as they were reported in annual rather than quarterly windows. Consequently, OOT was not feasible for the AFS due to limited access to data for model training. Furthermore, the limited code space in the AFS also reduced its statistical power and the model’s sensitivity to the characteristics of its individual codes.

Finally, the study did not benchmark FeePredict against alternative model families such as gradient boosting, support vector machines, or sequence models like Hidden Markov Models. Random Forest was selected for the reasons outlined in Section 2.4, but a formal head-to-head comparison would strengthen the case for the chosen architecture and is a natural extension of this work.

## 6 Conclusion

FeePredict demonstrates that changes to Medicare reimbursement rates are not random but are predictable based on publicly available information about the Medicare fee schedule. The fee schedule pipelines achieved concordance indices of 0.815–0.998 and reduced the mean absolute error of change-row rates by 29%–85% compared to a baseline model that assumed zero changes to the reimbursement rates for the CLFS, PFS, and DMEPOS codes; all results were statistically significant (*p <* 0.001), and label shuffle tests for model leakage confirmed the absence of such leakage. Furthermore, out-of-time validation indicates that the model maintains its predictive power for the CLFS (OOT C-Index = 0.854) and DMEPOS (OOT C-Index = 0.972) reimbursement rates, but begins to degrade for the PFS codes during the period after training data collection; this indicates that while FeePredict is generally applicable to the population of codes within the CLFS and DMEPOS, it may not be generalizable to the PFS codes in the future. The level of predictability of each reimbursement rate schedule correlates with the determinism of its rate changes.

Future work in this area may involve creating an annual update for the PFS code pipeline following the announcement of the conversion factor that Medicare will apply to all payment rates each year; these conversion factors are published well in advance of their implementation in July of each year. Additionally, extending the pipeline to the Medicaid and commercial payer rate schedules, which use the Medicare rates as a basis for calculation, is also a logical future development for the FeePredict system. Finally, expanding the observation window beyond 2026 as data becomes available will allow the model to incorporate additional data from the fee schedule, enabling FeePredict to better assess generalizability beyond the training data.

Overall, the development of FeePredict is beneficial to those organizations that are reimbursed according to the Medicare fee schedule; whether such organizations are health systems, laboratories, ambulances, or suppliers of durable medical equipment, FeePredict will allow them to better anticipate changes to their reimbursements prior to the Medicare updates being published each year for proactive financial planning.

## Data Availability

All fee schedule data are publicly available from the CMS website (https://www.cms.gov). Code and processed datasets will be made available upon publication at https://github.com/AI4Path-Lab/FeePredict.

## Ethics Statement

All data used in this study are publicly available and were downloaded directly from the CMS website (cms.gov). No patient-level data, protected health information, or insurance claims data were accessed or used at any stage. Institutional Review Board (IRB) approval was not required.

## Author Contributions

Ishan Patel conceptualized the modeling framework, developed the data processing and machine learning models, conducted the experiments and statistical analyses, and wrote the manuscript. Alex Leyva contributed to the conceptualization of the methodology to be used in the study. Khalid Niazi supervised the study, provided feedback on the manuscript, and edited the manuscript.

## Data Availability

All fee schedule data are publicly available from the CMS website (https://www.cms.gov). Code and processed datasets will be made available upon publication at [repository link].

## Supplementary Material

**Figure S1:**
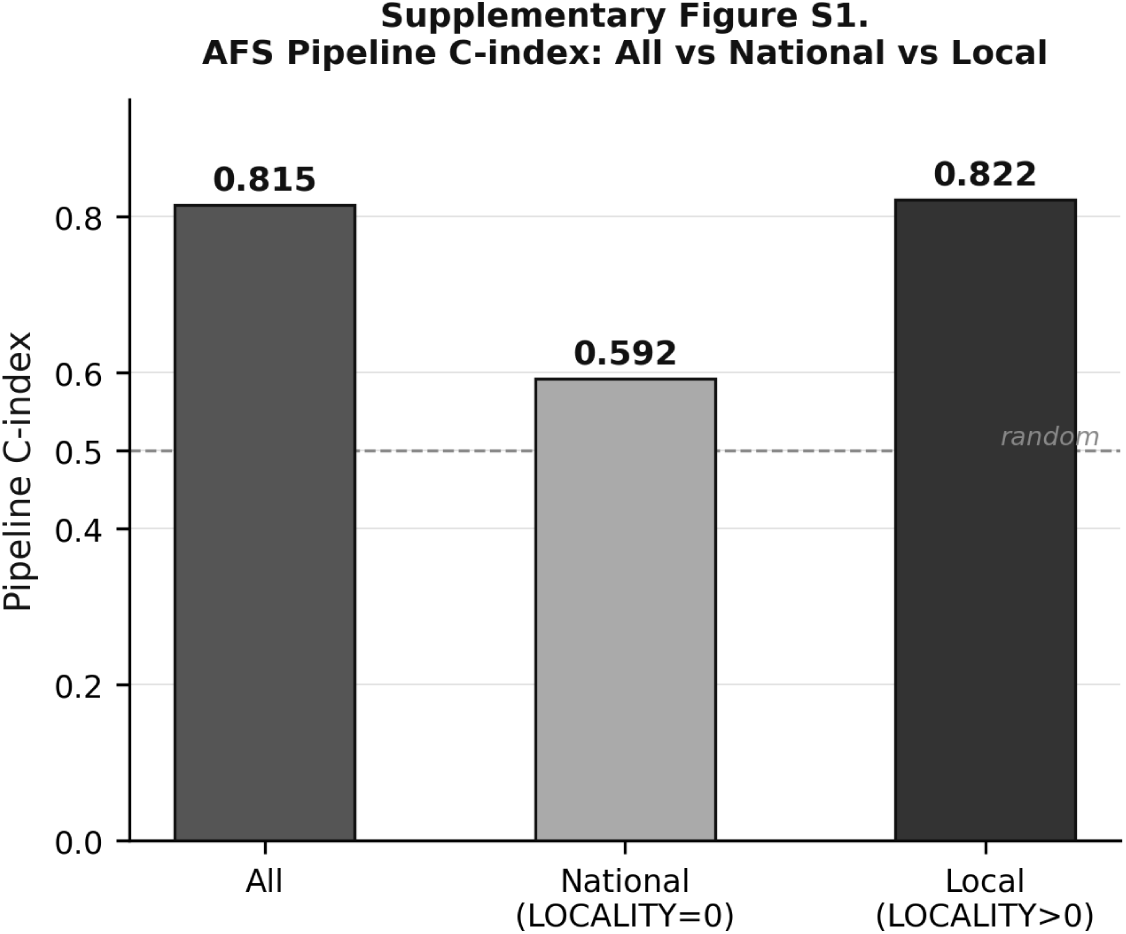
AFS pipeline C-index stratified by locality subgroup. “All” reflects the overall LOGO-CV C-index (0.815). “National” (LOCALITY = 0) reflects performance on codes with no locality adjustment (C-index = 0.592). “Local” (LOCALITY *>* 0) reflects performance on codes with locality-specific GPCI adjustments (C-index = 0.822). The dashed line indicates chance performance (C-index = 0.5).

